# Spatiotemporal dynamics and environmental drivers of listeriosis hospitalizations in Spain (2016–2024): A One Health perspective

**DOI:** 10.64898/2026.09.15.26363098

**Authors:** Rafael Garcia-Carretero, Beatriz Valle-Borrego, Clara Peiro-Villalba, Carmen de-Juan-Alvarez, Gema Cenzual-Alvarez, Sara-Maria Quevedo-Soriano

## Abstract

**Background:** *Listeria monocytogenes* causes invasive infections with high mortality. However, ecological studies linking human incidence to livestock and weather remain scarce. This retrospective study analyzed the geographical distribution of human listeriosis in Spain using nationwide hospitalization data, livestock censuses, and meteorological records.

**Methods:** Negative binomial regression models, spatial autocorrelation (Moran’s Index), and bivariate local indicators of spatial association (LISA) analysis were applied.

**Results:** Between 2016 and 2024, 4,391 hospital admissions were recorded. The national mean incidence was 1.03 per 100,000 inhabitants, peaking in 2019. Bovine density was the primary environmental driver, with an 11% increased risk per 10 heads/km^2^. Weather–livestock synergies were observed: bovine-associated risk increased exponentially at higher temperatures (*∼*21°C) and lower humidity (53%), whereas caprine-associated incidence peaked at cooler temperatures (*∼*13°C). Spatial mapping revealed northern clusters for bovine reservoirs and southern segregation for caprine reservoirs, with no associations for ovine or porcine livestock.

**Conclusions:** Zoonotic risk is environmentally conditioned, with weather acting as a biological trigger. This study highlights the critical role of ruminant farming practices and the need for a “One Health” surveillance approach that integrates meteorological and farming data to predict seasonal outbreaks and optimize food chain safety.

## Introduction

*Listeria monocytogenes* is a Gram-positive bacterium that causes the invasive disease listeriosis [1]. Human clinical syndromes are uncommon, occurring mainly in immunosuppressed individuals, newborns, the elderly, pregnant women, and occasionally healthy patients, in whom *L. monocytogenes* may cause self-limited febrile gastroenteritis [2–4].

Although infrequent, invasive infection can produce severe syndromes such as neuroinvasive disease and bacteremia. Listeriosis during pregnancy may result in spontaneous abortion, premature delivery, or neonatal infection [3, 4]. Outside of the pregnancy and neonatal period, immunosuppression, malignancies, and chronic conditions such as liver cirrhosis and chronic kidney disease are the main underlying, predisposing factors. Sepsis due to listeriosis can also occur in immunosuppressed adults and the elderly, often presenting as fever without a clear focus. The most severe clinical manifestation is neurological involvement, including meningoencephalitis, rhombencephalitis, or brain abscesses [3–9].

Despite its relatively low incidence compared to other foodborne diseases, listeriosis poses a major global public health challenge because of its high hospitalization and mortality rates. In the European Union, its incidence has shown an upward trend over the past decade, highlighting the urgent need to better understand the transmission dynamics of this pathogen [10, 11].

*L. monocytogenes* is also a frequent pathogen in veterinary medicine, causing neurological diseases in ruminants. As it can contaminate unpasteurized dairy or raw foods, it is considered one of the most virulent foodborne pathogens [2, 11].

Unlike many foodborne pathogens, *L. monocytogenes* is ubiquitous and resilient, capable of surviving in adverse environmental conditions, including moist soils, stagnant water, and extreme temperatures [12]. Its zoonotic nature is well documented, with bovine livestock serving as a primary reservoir [13, 14]. Fecal excretion of *Listeria* is significantly higher in cattle than in other ruminants, often linked to consumption of poor-quality or improperly fermented silage [15, 16].

Nevertheless, ecological studies analyzing the association between human disease incidence and non-clinical factors such as environmental conditions and livestock density at a national scale remain scarce. Human cases are typically linked to contaminated ready-to-eat products rather than direct environmental or occupational exposure [17]. In Spain, however, the availability of detailed administrative and geographical data enables ecological analyses of how regional livestock distribution and weather conditions influence disease incidence. Because listeriosis is not consistently subject to exhaustive epidemiological surveillance across all healthcare levels, hospitalization registries provide a robust and reliable estimate of incidence and burden. Spain’s nationwide hospitalization registries are compulsory and have proven to be an ideal source for nationwide ecological research [18].

Our main objective was to analyze the geographical distribution of hospitalized infections caused by *L. monocytogenes* in Spain between 2016 and 2024 to identify the relationships between this distribution and external factors (i.e., livestock density and meteorological factors). To this end, we used advanced regression models and spatial analysis techniques. The secondary objective was to identify critical control areas where listeriosis converges with specific weather conditions, through analysis of those relationships.

## Methods

### Study design and geographical and temporal scope

We conducted a nationwide, retrospective ecological study to describe the geographical distribution of patients hospitalized with *L. monocytogenes* infections in Spain between 2016 and 2024. Specifically, we examined the associations between infection incidence and environmental exposures at the population level. Aggregate data included hospitalization rates across Spanish regions, livestock activity, and weather conditions. Analyses were performed at the regional and provincial levels, covering a population of 48,619,695 (as of January 1, 2024), with minor annual variation. Spain’s administrative structure consists of 17 autonomous communities (first-level divisions) and 2 autonomous cities. The 17 autonomous communities are further subdivided into 50 provinces.

### Data collection

#### Incidence of listeriosis

Hospitalization data were obtained from the Minimum Basic Data System at Hospitalization (MBDS-H), a compulsory, standardized nationwide registry based on discharge reports from both public and private hospitals. Nearly 95% of hospitals are covered and about 97% of all discharge reports are included in this database from the Spanish National Health System [19]. We identified 4,394 admissions between 2016 and 2024 with a diagnosis of listeriosis, coded using the 10th Clinical Revision of the International Classification of Diseases (ICD-10-CM) diagnoses for listeriosis (code A32). Because MBDS-H data are released with a 1-year delay, the Spanish Ministry of Health provided updated records through December 31, 2024. We summarized case counts at regional and provincial levels and calculated hospitalization rates per 100,000 population.

#### Livestock data

Livestock census data were obtained from the National Statistics Institute (*Instituto Nacional de Estadística*, INE) [20]. We collected data on bovine, caprine, ovine, and porcine population densities (heads/km^2^), calculated using provincial land area. The most recent census data available were from 2023. Assuming minimal changes between 2023 and 2024, we used the 2023 data.

#### Weather conditions

Meteorological data were obtained from the Spanish National Agency for Meteorology (*Agencia Española de Meteorología*, AEMET) [21]. Regional and provincial mean annual values for 2024 were calculated from daily records, including mean temperature (ºC), mean accumulated precipitation (mm), and humidity (%).

#### Statistical Analysis

Because observed values were aggregated at both regional and provincial levels, we applied lattice data analysis (spatial analysis) [22]. This approach interprets information linked to physical locations to identify patterns, trends, and territorial relationships. Our objective was to transform raw geographical data into insights that clarify spatial associations. Spatial relationships were managed using Geographic Information Systems (GIS) and open data sources [23].

Our analyses followed four steps: (a) fit the regression model; (b) test for spatial correlation (spatial dependence); (c) incorporate significant spatial correlations into the model; and (d) identify critical areas of convergence between high listeriosis incidence and high livestock density.

To fit the regression model, we used negative binomial (NB) regression, which is appropriate for count data with over-dispersion. Over-dispersion occurs when variance exceeds the mean, violating the Poisson regression’s assumption of equal mean and variance. NB regression generalizes the Poisson model by adding a dispersion parameter to account for extra variability, yielding more reliable estimates. Our model produced incidence rate ratios (IRRs), representing the expected change in counts when a predictor variable changes by one unit.

Because geographically proximate data points are often more correlated than distant ones, we accounted for spatial autocorrelation, which occurs when events in one region are influenced by neighboring regions. Risk of listeriosis was decomposed into covariates (livestock variables), weather conditions, neighborhood effects, and random heterogeneity. To capture spatial dependence, we used Moran’s Index (Moran’s I), which measures whether cases are randomly distributed or geographically clustered. It was used to guide the choice of spatial model (in our case, geographically weighted NB regression).

It was calculated on model residuals to evaluate whether errors were spatially grouped. Neighbor lists were constructed based on contiguous boundaries (e.g., Galicia is adjacent to Asturias but not to Murcia).

In sum, we combined NB regression with Moran’s I to account for spatial dependence. A row-standardized spatial weights matrix was constructed using the Queen’s contiguity criterion to define neighborhood relationships among the 50 Spanish provinces.

#### Model comparison

We computed multiple linear regression models with both main and interaction effects, using NB regression to achieve mathematical convergence. We produced three models. The first was an unadjusted model, using densities of livestock in heads/km^2^. The second was a weather-adjusted model, which added weather conditions to the previous model. These models analyzed main effects, measuring the independent impact of a single variable on count data while assuming other variables remained constant. That is, livestock and weather conditions were considered independent predictor variables. The third was an interaction model, which included those individual effects plus interaction terms, representing whether the combined influence of two variables created an effect greater or less than their sum, i.e., synergy. This interaction model tested whether the effect of livestock (*x_1_*) on the count data changed depending on the level of weather conditions (*x_2_*). Assuming non-linearity of interactions among covariates, we ran NB regression with an interaction model (instead of an additive model) to examine the effect by adding a product term (*x_1_* × *x_2_*) to the regression equation. Synergy was mathematically captured by the product term, revealing non-parallel slopes and a more complex, non-additive relationship. If regression coefficients were positive and significant (p *<* 0.05), the model would demonstrate that risk for listeriosis increased non-linearly when densities of livestock and certain weather conditions concurred. This would suggest, for example, that the effect of having large numbers of ruminants is more dangerous in a region with certain weather conditions than in one without those conditions.

#### Identification of critical control areas

Finally, using a spatial convergence approach, we identified critical control areas. To achieve spatial convergence we used bivariate local indicators of spatial association (LISA). This geographic analysis technique examines the spatial relationship between two different variables, identifying areas where they cluster together (hotspots/coldspots) or where one is significantly associated with its neighbors’ values. It helps uncover localized patterns, revealing complex interdependencies between phenomena such as health indicators and environment, often visualized in maps showing significant positive or negative spatial correlations. In our context, LISA would identify which regions are hotspots. By analyzing livestock density and case rate, it classifies regions as High-High (high livestock density and high case rates, i.e., hotspots) and Low-Low (coldspots or low-risk areas). The significance of local spatial clusters was assessed using 999 random permutations, with a pseudo p-value threshold *<* 0.05 to identify statistically significant hotspots and outliers. LISA does not prove causation; rather, it identifies significant spatial convergence.

Of note, to compute bivariate LISA we used hospitalization rate per 100,000 population instead of absolute count. The rationale for this decision was the need to normalize the burden of disease relative to the resident population in each spatial unit. This avoided bias related to provinces with higher demographic density and allowed the analyses to identify areas of spatial convergence between livestock and population-attributable disease risk. Thus, our approach can be considered methodologically robust.

A p-value *<* 0.05 was considered statistically significant. In the case of Moran’s I, p *>* 0.05 indicated that residuals were randomly distributed, the NB regression was sufficient, and no more spatial models should be used. All analyses were conducted using R version 4.5.2. Packages such as spdep, MASS, rgeoda, ggplot2, and sf were used for the analyses.

#### Ethical Considerations

The study was conducted in compliance with the Declaration of Helsinki. Data were obtained from open sources provided by the Spanish Ministry of Health [24], AEMET [21], and INE [20]. Hospitalization data were anonymized and deidentified, ensuring confidentiality and privacy. Because no names or personal information were recorded, no additional patient consent was required.

## Results

In total, 4,391 hospital admissions with a diagnosis of listeriosis were recorded in Spain between 2016 and 2024. The overall national hospitalization rate during the study period was 1.03 per 100,000 population. Temporal analysis showed a peak in 2019, with 750 admissions and a rate of 1.59 per 100,000. This was followed by a sharp decline in 2020 (371 cases; 0.78 per 100,000) and stabilization between 0.99 and 1.0 per 100,000 during 2022–2024 (Supplementary Table 1). Supplementary Figure 1 shows monthly case trends, with more cases observed between May and September.

Geographically, at the regional level, the highest absolute number of admissions occurred in Catalunya (n=992), Andalucía (n=792), and Madrid (n=722), as shown in Supplementary Table 1. However, the highest mean hospitalization rates were observed in Cantabria (1.62), Catalunya (1.43), Euskadi (1.34), Galicia (1.3), and La Rioja (1.22). No cases were recorded in the autonomous cities of Ceuta and Melilla during the study period.

At the provincial level, the analyses revealed a heterogeneous distribution (Figure 1). Valladolid, Lugo, and Seville had the highest hospitalization rates (Supplementary Table 2). Overall, northwestern provinces were high-incidence areas, unlike central regions such as Extremadura and Castilla-La Mancha.

**Figure 1.**
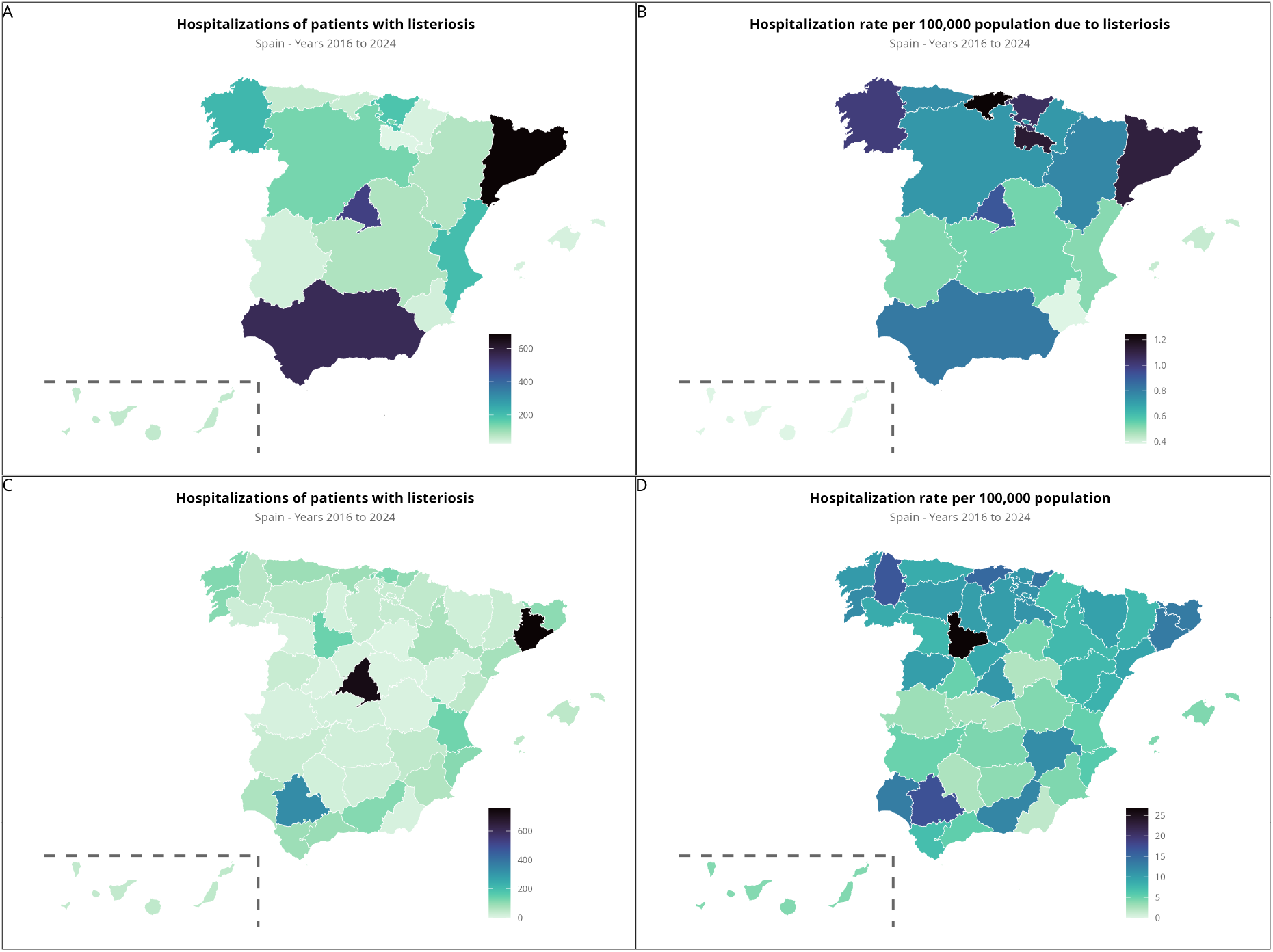
Geographical distribution of listeriosis cases in Spain at the regional level (A and B) and provincial level (C and D)

Livestock, in terms of farms and density by province, are summarized in Supplementary Figures 2 and 3. Bovine livestock predominates in northern Spain, while caprine livestock does in southern regions.

Regarding weather conditions, the central-southern provinces showed the highest temperatures, while the greatest humidity and precipitation occurred in the northern regions (Supplementary Figure 4).

### Spatial regression

In NB regression analysis, the initial, unadjusted model identified bovine livestock as the primary driver of listeriosis (Table 1). It showed an 11% increased risk in IRR for every 10 heads/km^2^ increase in bovine density (IRR = 1.11, p = 0.003). In contrast, no significant associations were found for ovine, caprine, or porcine densities. The weather-adjusted model also did not identify effects of livestock or weather on listeriosis incidence. However, the interaction model revealed significant relationships. Specifically, it indicated a synergy between bovine livestock and temperature (IRR = 1.09, p = 0.008), and between caprine livestock and temperature (IRR = 1.303, p *<* 0.001), indicating that the impact of livestock reservoirs on case rates increases with high temperatures. By contrast, interactions between bovine and caprine livestock, on the one hand, and humidity on the other, showed a protective effect. Figure 2 presents the results of the interaction model for bovine and caprine livestock with humidity and temperature.

**Table 1.** Negative binomial regression model based on density (10 heads/km^2^) at the region level.

|  | Unadjusted regression model |  |  | Adjusted regression model |  |  | Adjusted regression model with interactions |  |  |
| --- | --- | --- | --- | --- | --- | --- | --- | --- | --- |
|  | IRR | 95% CI | P-value | IRR | 95% CI | P-value | IRR | 95% CI | P-value |
| Bovine density (10 heads/km<sup>2</sup>) | 1.114 | 1.005–1.238 | 0.033 | 1.052 | 0.924–1.200 | 0.458 | 2.151 | 0.37–12.871 | 0.398 |
| Ovine density (10 heads/km<sup>2</sup>) | 0.937 | 0.886–0.993 | 0.022 | 0.944 | 0.895–0.996 | 0.034 | 0.877 | 0.852–0.901 | <0.001 |
| Caprine density (10 heads/km<sup>2</sup>) | 0.782 | 0.646–0.953 | 0.01 | 0.992 | 0.755–1.295 | 0.948 | 0.007 | 0–0.113 | <0.001 |
| Porcine density (10 heads/km<sup>2</sup>) | 1.003 | 0.988–1.020 | 0.701 | 1.011 | 0.994–1.029 | 0.228 |  |  |  |
| Humidity (%) |  |  |  | 1 | 0.968–1.034 | 0.096 | 1.046 | 1.017–1.077 | 0.002 |
| Temperature (°C) |  |  |  | 0.929 | 0.851–1.015 | 0.983 | 0.799 | 0.724–0.883 | <0.001 |
| Precipitation (mm) |  |  |  | 1 | 1–1.001 | 0.381 | 0.999 | 0.998–0.999 | <0.001 |
| Bovine density × Temperature<sup>1</sup> |  |  |  |  |  |  | 1.09 | 1.023–1.161 | 0.008 |
| Bovine density × Humidity<sup>1</sup> |  |  |  |  |  |  | 0.966 | 0.952–0.981 | <0.001 |
| Caprine density × Temperature<sup>1</sup> |  |  |  |  |  |  | 1.303 | 1.157–1.467 | <0.001 |
| Caprine density × Humidity<sup>1</sup> |  |  |  |  |  |  | 0.981 | 0.963–0.999 | 0.042 |
|  | Moran I statistic = -0.049,<br>standard deviation = 0.137,<br>p-value = 0.445 |  |  | Moran I statistic = -0.128,<br>standard deviation = -0.354,<br>p-value = 0.638 |  |  | Moran I statistic = -0.092,<br>standard deviation = -0.144,<br>p-value = 0.557 |  |  |

<sup>1</sup>Interaction model. IRR: incidence rate ratio, CI: confidence interval. The porcine density variable was dropped from the final model.

**Figure 2.**
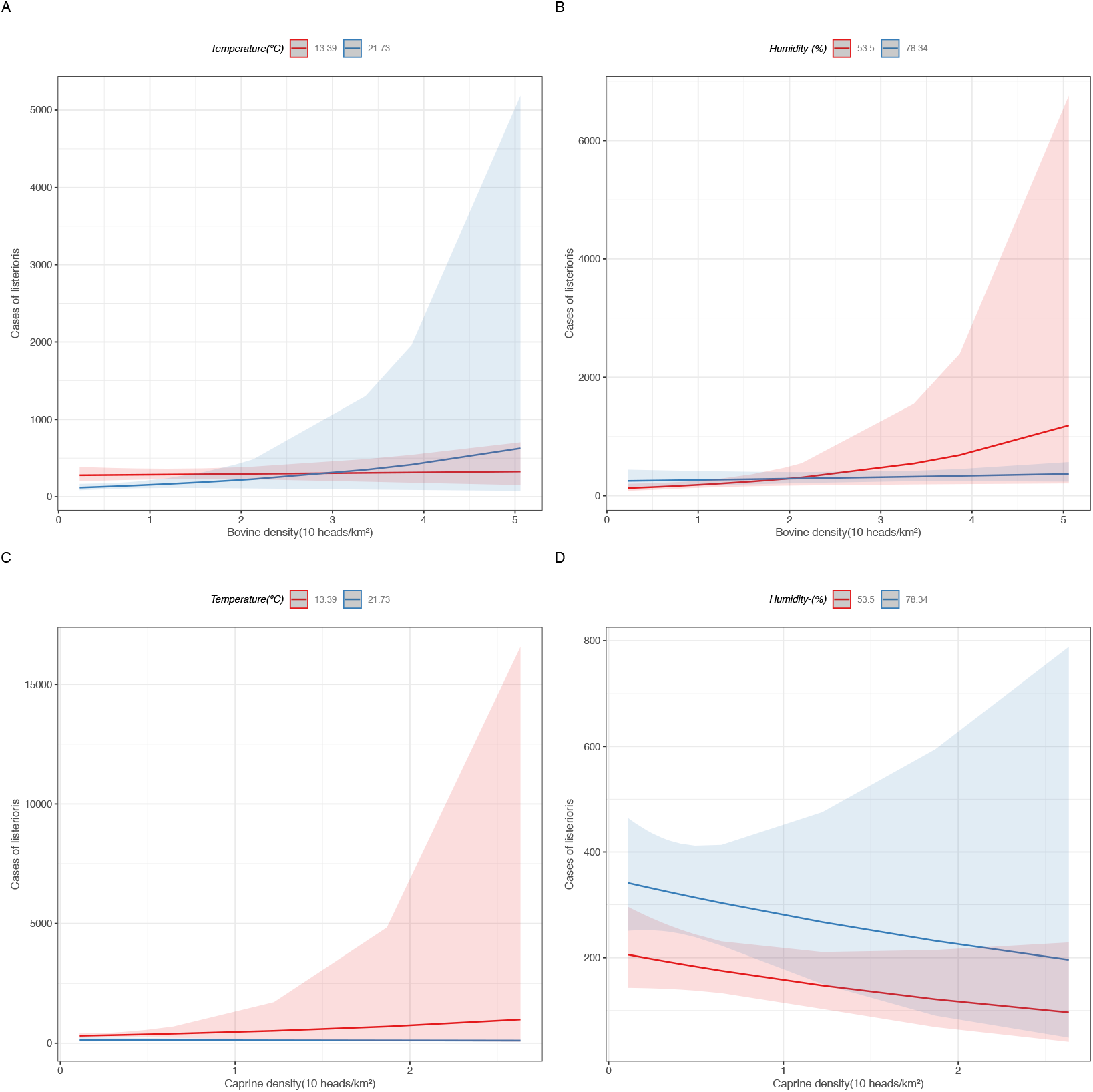
Multiple linear regression with interaction effects and 95% confidence intervals.

Moran’s I was calculated on the residuals of the adjusted models, yielding a value of –0.09 (p = 0.56) for the interaction model. These nonsignificant p-values indicate that the models successfully accounted for spatial dependency, so further spatial modeling was not required. Model parsimony and goodness-of-fit were also assessed using the Akaike Information Criterion (not shown).

### Analysis of marginal effects of interaction terms

Figure 2 shows effect plots of the interaction terms in the regression model, illustrating the impact of livestock density on the case rate of listerioris. Case counts vary as weather conditions change. Each panel in Figure 2 presents predicted values across selected predictors, i.e., livestock and weather-related variables. For instance, panel A shows predicted case rates as heads/km^2^ vary at two temperatures (21°C and 13ºC). The red and blue lines are not parallel, indicating that temperature modifies the effect of livestock density on case rates. At higher temperatures, an increase in density leads to an exponential increase in disease incidence, whereas the slope is less steep at lower temperatures. Analyses of marginal effects confirmed that the risk associated with bovine density is temperature-dependent: at *∼*21°C, listeriosis rates increase exponentially with bovine density, while the relationship is absent at *∼*13°C. This pattern suggests that higher temperatures act as a trigger, amplifying the risk of listeriosis associated with bovine livestock.

A similar phenomenon was observed for caprine livestock (Figure 2, panel C), but in the opposite direction: case rates increase at lower temperatures, with no changes at higher temperatures. Thus, for caprine livestock, cooler temperatures act as the trigger of incidence. Although statistically significant, the effect of caprine density in relation to temperature is less pronounced.

Regarding humidity and bovine livestock (Figure 2, panel B), low humidity (53%) is the trigger. Conversely, as shown in panel D, there is a negative interaction between caprine livestock and humidity. Unlike the synergies observed in bovine models, this analysis revealed a decrease in incidence with an increase in caprine density in high-humidity environments. Although counterintuitive, this may reflect differences in livestock management or ecological competition between reservoirs. The latter could suggest that caprine livestock, under certain conditions, functions not as a modifier but as an effect-size attenuation factor.

### Bivariate LISA Analysis

Finally, analysis of spatial autocorrelation using bivariate LISA revealed heterogeneous patterns and territorial disparities regarding the association between livestock density and listeriosis (Figure 3). For bovine density (panels A and B), both regional and provincial analyses identified significant High-High clusters (representing critical control areas where high density converges with high listeriosis incidence), concentrated almost exclusively in northern Spain (Asturias). In contrast, central and southern Spain showed low convergence (Low-Low or Low-High). Interestingly, when analyzed at the provincial level, Toledo was identified as a High-Low hotspot, suggesting a dilution effect. Caprine density (panels C and D) showed the geographically opposite pattern. Clusters of convergence (High-Low or High-High) were identified in southern Spain. A *post-hoc* Kruskal**–**Wallis test confirmed that these regional differences were not random. Significant p-values (*<*0.003 in all cases) supported the robustness of the observed disparities.

**Figure 3.**
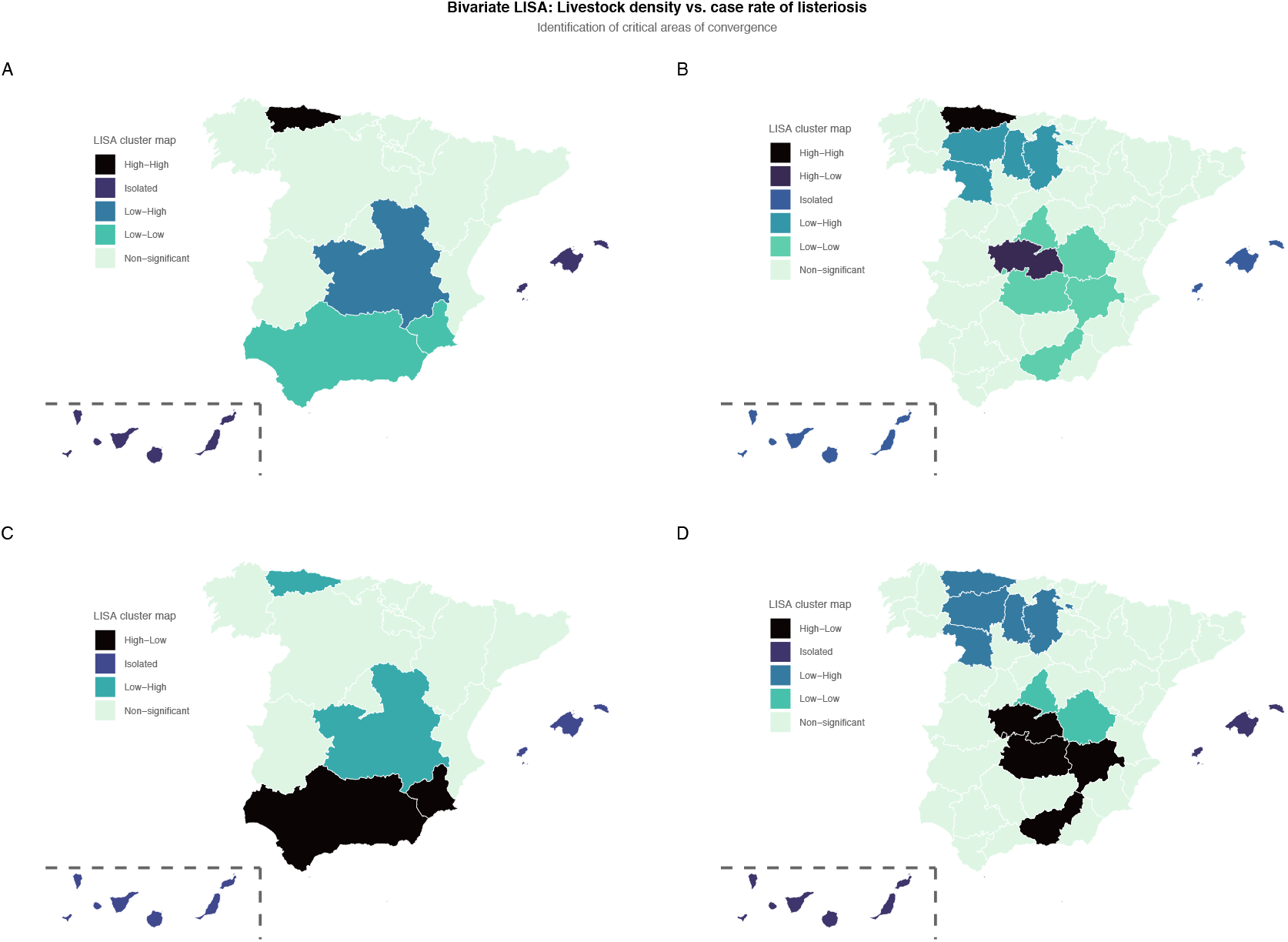
Bivariate LISA cluster maps showing hotspots and coldspots in Spain for bovine livestock (A, B) and caprine livestock (C, D) at the regional and provincial levels, respectively.

## Discussion

### Overall results

This nationwide study demonstrates correlations between hospitalizations due to *L. monocytogenes* and environmental factors in Spain, particularly weather conditions and livestock density. Between 2016 and 2024, the mean hospitalization rate was 1.03 per 100,000 population, with 0.99 in 2024. Surveillance of listeriosis is mandatory in almost all countries in the European Union (EU), and coverage in Spain is estimated at 97% [11]. In 2024, the EU notification rate was 0.69 per 100,000 population, the highest since 2007, with Luxembourg (1.2), Sweden (1.1), and Denmark (1.1) reporting the highest rates [11]. At the country level, the European Food Safety Authority (EFSA) and the European Centre for Disease Prevention and Control (ECDC) noted significant increasing trends in several member states, including Spain. Most reported cases (99.3%) were acquired within the EU, and nearly all required hospitalization, supporting the robustness of our descriptive analysis.

### Bovine and caprine livestock as predominant reservoirs

Our analysis highlights region-specific associations across Spain, aligning with the established role of livestock as primary reservoir of listeriosis [25]. While *Listeria* can infect a wide range of domestic mammals, prevalence studies indicate that cows, sheep, and goats carry higher risk compared to pigs [14]. Several factors may underlie these associations.

For example, poorly maintained or improperly fermented silage can facilitate *Listeria* proliferation [15,16], creating persistent environmental pressure and increasing the likelihood of contamination in the food chain [26]. However, transmission dynamics between the environment, ruminants, and humans remain incompletely understood [27–29]. Given the role of domestic ruminants as reservoirs of pathogenic strains, further work is needed to characterize strain virulence and pathways of human infection. Regarding cattle, Moura et al. [30] reported a high prevalence of hyper-virulent strains in bovine livestock.

### Interaction regression modeling: synergistic effects of weather and livestock density

A key contribution of our study is the identification of a significant interaction between livestock density and weather-driven conditions. Our analysis of marginal effects reveals that the effect of livestock on the case rate depends on weather. Neither bovine nor caprine livestock act as stand-alone risk factors; rather, their impact is temperature- and humidity-dependent. Our interaction models demonstrate that the relationship is non-linear and dependent on regional weather conditions.

The results of the interaction model using NB regression showed improvement over the previous models (both unadjusted and weather-adjusted) when interaction terms were included. These models demonstrate that the interaction between livestock and weather conditions is non-additive but synergistic. Our findings suggest that the combination of heat and low humidity, on the one hand, and high-density livestock areas on the other, acts as a biological trigger, allowing clusters of active transmission or “hotspots.” We found exponential relationships indicating that temperature enhances transmission and/or environmental proliferation of the bacteria in areas of high livestock density.

At lower temperatures (*∼*13°C), caprine livestock was associated with an increase in cases, whereas at higher temperatures (*∼*21°C), bovine livestock was related to an increase. The interaction model also suggested that humidity may influence *Listeria* transmission synergistically with livestock. As humidity decreases (to *∼*53%), the risk increases exponentially as bovine livestock density increases.

The interaction model can also explain the observed seasonal spikes in listeriosis cases during the warmer months. The environmental abundance of *L. monocytogenes* is higher during summer, beginning in May (as seen in Supplementary Figure 1), probably related to the natural breeding season of cattle. High temperatures enhance the environmental proliferation and survival of *Listeria* in soil, silage, and water sources near high-density farming areas. Watanabe et al. [31] also reported an increase in the rate of listeriosis cases during the warm season (May to October). In contrast to our findings, another study conducted in Spain reported higher *Listeria* prevalence in bovine livestock during winter [32], likely linked to second lactation and associated fecal shedding.

Our interaction models explain observed data better than either unadjusted or adjusted models. However, our findings on the interaction between livestock and humidity, showing an attenuating effect, suggest complex dynamics in the environmental persistence of the pathogen that are not explained by our marginal effects analyses.

### Bivariate LISA and identification of critical control areas

The rationale for using bivariate LISA was to provide a complementary analysis beyond global regression models. Our NB regression results assume homogeneity among spatial relationships, yet in environmental epidemiology the relationship between a reservoir (livestock density) and an outcome (listeriosis case rate) can be spatially heterogeneous.

Our bivariate LISA maps add a spatial perspective to the regression models. The analysis of spatial clusters confirmed the existence of preferred ecological niches, with listeriosis risk in Spain showing marked heterogeneity and competitive displacement between reservoirs. Bovine livestock frequently serve as reservoirs of *Listeria*, shedding bacteria through feces. Competitive strain dominance may also play a role, with specific clonal complexes prevalent in livestock environments displacing minor strains under seasonal and regional pressures [32]. Identification of High-High convergence areas in northern regions aligns with Spanish zones where humidity, temperature, and precipitation facilitate poor ensilage preparation, storage, and/or consumption, known risk factors for Listeria infection among ruminants [26]. Thus, Asturias represents a geographical convergence of high bovine pressure and a favorable ecological niche characterized by high humidity and moderate-to-high temperatures. As discussed previously, this suggests that epidemiological risk depends not only on reservoir presence (bovine livestock in this case) but also on the geographical convergence of environmental conditions that favor bacterial persistence in the environment.

Caprine livestock also showed spatial segregation in southern regions. A phenomenon of territorial exclusion emerged: regions with high densities of bovine livestock lacked high densities of caprine livestock, and vice versa. This pattern suggests displacement and interspecific competition in farming areas. It may explain the negative or mitigating effect observed when computing both adjusted and unadjusted regression models for caprine livestock. Our interaction models, as discussed, indicate that synergy between bovine livestock and higher temperatures is critical in northern regions, while interactions between caprine livestock and lower temperatures increase risk in southern regions.

Interestingly, these distinct clustering densities may simply reflect regional differences in livestock distribution rather than disease-related mechanisms. The direct relevance to disease risk remains uncertain and requires further research.

The identification of such hotspots using bivariate LISA, combined with our interaction models, provides a basis for targeted public health and epidemiological surveillance measures addressing food chain safety and animal health in specific livestock farming territories across Spain.

### Bovine livestock and other livestock categories

We observed a strong association between human cases of listeriosis and bovine and caprine livestock, measured as density of heads per km^2^. From both epidemiological and veterinary perspectives, these findings are consistent with other studies on reservoirs of *L. monocytogenes*. The main source of infection for cattle is low-quality silage or poorly maintained fermented roughage [33]. Cows are fed large amounts of this silage, facilitating the infection cycle and widespread environmental contamination [26, 34]. The relationship between density (heads/hm^2^) and incidence rate suggests an effect of environmental pressure, as noted by Esteban et al. [25]. High densities of bovine farms generate large amounts of manure that must be managed. Cattle exposed to contaminated silage amplify transmission by increasing fecal shedding, thereby contributing to the maintenance and dispersal of *Listeria* in the environment [27, 33]. A study of 100 dairy farms in Portugal covering the 2020–2021 period reported prevalence rates of 8.3% in water, 12.5% in feces, and 12.0% in feed [35].

Because *Listeria* is highly resistant in water and soil, greater bovine density increases the likelihood of contamination reaching the food chain through vegetables or cross-contamination in processing plants, as demonstrated by Nightingale et al. [27].

We did not find a significant association between listeriosis cases and the density of ovine or porcine livestock, likely because they are less frequently linked to human epidemiological outbreaks [25]. Although *Listeria* can infect any ruminant, bovine, ovine, and caprine livestock are the primary reservoirs [25]. Studies of cattle show greater fecal excretion of *Listeria* compared to pigs and other small ruminants [25, 27]. In Spain, ovine and caprine livestock are typically raised under extensive or semi-extensive systems, whereas bovine livestock is managed intensively. Sheep and goats are therefore more scattered across the landscape, reducing pathogen burden per km^2^ compared to intensive bovine or caprine systems that involve silage and manure [29]. Moreover, most food products associated with listeriosis outbreaks in Spain are linked to bovine and beef products, such as raw-milk cheeses [36].

### One Health approach and sustainability

Our results highlight the need for a “One Health” approach to listeriosis prevention. Incorporating meteorological data and livestock distribution into public health surveillance systems could improve the prediction of seasonal outbreaks and facilitate the implementation of more effective food safety controls in high-risk regions.

### Conclusions

This nationwide ecological study provides robust evidence that bovine and caprine livestock density are primary environmental drivers of human listeriosis hospitalizations in Spain. Our findings go beyond simple associations by demonstrating critical synergistic interactions. The incidence of listeriosis increases exponentially when high-density livestock converges with specific weather variables. This suggests that zoonotic risk should not be interpreted as additive but rather as an environmentally conditioned phenomenon. Risk is triggered by bovine density at high temperatures and by caprine density at low temperatures, likely enhancing the environmental proliferation of *L. monocytogenes* and its transmission from ruminant reservoirs to the human population.

The integration of spatial analysis and interaction models using bivariate LISA allowed us to identify local clusters of spatial dependence, or critical control areas. These hotspots represent priority zones for targeted environmental and veterinary surveillance. Our results provide an empirical basis for designing control strategies focused on high-convergence zones. The study offers more than statistical analysis; it provides a practical tool for surveillance interventions in public health aimed at reducing disease incidence.

The absence of significant associations with ovine or porcine livestock suggests that preventive efforts should remain focused on the bovine and caprine supply chain and the management of intensive livestock farming environments.

## Data Availability

The database was built from open data, and the original
datasets can be separately downloaded from their open sources.
However, we have also uploaded the compiled, final dataset
that supports the findings of this study to a public reposi-
tory, available at https://github.com/rafalinux/
listeria.

https://github.com/rafalinux/listeria

## Declarations

### Authors’ contributions

Dr. Garcia-Carretero conceived and designed the study, wrote the first draft of the manuscript, and preprocessed and analyzed the data. Dr. Valle-Borrego, Dr. Peiro-Villalba, Dr. de-Juan-Alvarez, and Dr. Cenzual-Alvarez made substantial contributions to the interpretation of the results, critically reviewed the first draft of the manuscript, made valuable suggestions, and contributed to the visualization of the data. Dr. Quevedo-Soriano supervised the project and critically reviewed and edited the final draft of the manuscript. All authors read and approved the final manuscript.

### Availability of data and materials

The database was built from open data, and the original datasets can be separately downloaded from their open sources. However, we have also uploaded the compiled, final dataset that supports the findings of this study to a public repository, available at https://github.com/rafalinux/listeria.

### Ethics approval and consent to participate

The study was conducted in compliance with the Declaration of Helsinki. Data were obtained through a formal request to the Spanish Ministry of Health. Patient records were provided anonymized and de-identified, ensuring confidentiality and privacy. Because no names or personal information were recorded, no additional patient consent was required.

### Consent for publication

Not applicable.

### Declaration of generative AI and AI-assisted technologies in the manuscript preparation process

During the preparation of this work the authors did not use AI-assisted technologies. The authors take full responsibility for the content of the published article.

