## Supplementary Material for "Spatiotemporal dynamics and environmental drivers of listeriosis hospitalizations in Spain (2016–2024): A One Health perspective"

#### This PDF file includes:

Supporting text
Figs. S1 to S4
Tables S1 to S2

### Supporting Information Text

### Methodology

To address potential multicollinearity among selected covariates, we evaluated correlations between variables. Multicollinearity introduces redundancy and can destabilize regression estimates. We quantified this risk using the variance inflation factor (VIF), which measures how much the variance of a regression coefficient is inflated by collinearity. Ideally, VIF values should be as low as possible. Variables with high VIF values were excluded from the final model. This approach produced simpler, more stable models without compromising accuracy. Following a conservative rule of thumb, we retained only variables with VIF $<5$ in the final multivariate analyses.

### Results

**Table S1. Geographical distribution of Listeria monocytogenes infections across different regions of Spain**

| Region | 2016 | 2017 | 2018 | 2019 | 2020 | 2021 | 2022 | 2023 | 2024 | Total |
|---|---|---|---|---|---|---|---|---|---|---|
| Andalucía | 57 (0.68) | 47 (0.56) | 78 (0.93) | 331 (3.93) | 71 (0.84) | 36 (0.42) | 54 (0.63) | 64 (0.76) | 54 (0.63) | 792 (1.04) |
| Aragón | 14 (1.07) | 4 (0.31) | 16 (1.22) | 13 (0.99) | 9 (0.68) | 21 (1.58) | 12 (0.9) | 10 (0.75) | 14 (1.04) | 113 (0.95) |
| Asturias | 17 (1.63) | 6 (0.58) | 12 (1.17) | 7 (0.68) | 6 (0.59) | 6 (0.59) | 12 (1.19) | 15 (1.49) | 8 (0.79) | 89 (0.97) |
| Illes Balears | 3 (0.27) | 5 (0.45) | 14 (1.24) | 3 (0.26) | 6 (0.51) | 4 (0.34) | 4 (0.34) | 7 (0.58) | 7 (0.57) | 53 (0.51) |
| Canarias | 8 (0.38) | 6 (0.28) | 15 (0.7) | 6 (0.28) | 13 (0.6) | 10 (0.46) | 14 (0.64) | 8 (0.36) | 13 (0.58) | 93 (0.48) |
| Cantabria | 6 (1.03) | 7 (1.21) | 11 (1.9) | 19 (3.27) | 10 (1.72) | 9 (1.54) | 8 (1.37) | 11 (1.87) | 4 (0.68) | 85 (1.62) |
| Castilla y León | 25 (1.02) | 27 (1.11) | 27 (1.12) | 29 (1.21) | 19 (0.79) | 25 (1.05) | 32 (1.35) | 37 (1.55) | 31 (1.3) | 252 (1.17) |
| Castilla - La Mancha | 15 (0.73) | 8 (0.39) | 17 (0.84) | 16 (0.79) | 9 (0.44) | 17 (0.83) | 10 (0.49) | 12 (0.58) | 22 (1.05) | 126 (0.68) |
| Catalunya | 126 (1.67) | 104 (1.38) | 99 (1.3) | 104 (1.36) | 91 (1.17) | 115 (1.48) | 106 (1.37) | 118 (1.49) | 129 (1.61) | 992 (1.43) |
| Comunitat Valenciana | 30 (0.6) | 34 (0.69) | 37 (0.75) | 34 (0.68) | 16 (0.32) | 37 (0.73) | 33 (0.65) | 33 (0.63) | 34 (0.64) | 288 (0.63) |
| Extremadura | 6 (0.55) | 6 (0.56) | 8 (0.75) | 10 (0.94) | 6 (0.56) | 9 (0.85) | 3 (0.28) | 6 (0.57) | 5 (0.47) | 59 (0.61) |
| Galicia | 30 (1.1) | 34 (1.26) | 26 (0.96) | 33 (1.22) | 24 (0.89) | 41 (1.52) | 40 (1.49) | 44 (1.63) | 44 (1.63) | 316 (1.3) |
| Madrid | 79 (1.22) | 68 (1.04) | 108 (1.64) | 100 (1.5) | 59 (0.87) | 71 (1.05) | 95 (1.41) | 68 (0.99) | 74 (1.06) | 722 (1.2) |
| Murcia | 8 (0.55) | 7 (0.48) | 4 (0.27) | 7 (0.47) | 5 (0.33) | 6 (0.4) | 12 (0.78) | 9 (0.58) | 8 (0.51) | 66 (0.49) |
| Nafarroa | 7 (1.09) | 8 (1.24) | 4 (0.62) | 5 (0.76) | 6 (0.91) | 4 (0.6) | 7 (1.05) | 3 (0.45) | 0 (0) | 44 (0.74) |
| Euskadi | 32 (1.46) | 24 (1.09) | 40 (1.82) | 26 (1.18) | 18 (0.81) | 33 (1.49) | 28 (1.27) | 33 (1.49) | 32 (1.44) | 266 (1.34) |
| La Rioja | 3 (0.95) | 4 (1.27) | 7 (2.22) | 7 (2.21) | 3 (0.94) | 3 (0.94) | 0 (0) | 4 (1.24) | 4 (1.23) | 35 (1.22) |
| Ceuta | 0 (0) | 0 (0) | 0 (0) | 0 (0) | 0 (0) | 0 (0) | 0 (0) | 0 (0) | 0 (0) | 0 (0) |
| Melilla | 0 (0) | 0 (0) | 0 (0) | 0 (0) | 0 (0) | 0 (0) | 0 (0) | 0 (0) | 0 (0) | 0 (0) |
| Total | 466 (1) | 399 (0.86) | 523 (1.12) | 750 (1.59) | 371 (0.78) | 447 (0.94) | 470 (0.99) | 482 (1) | 483 (0.99) | 4391 (1.03) |

Data are expressed in absolute values (hospitalization rate per 100,000 population).

#### Table S2. Listeriosis cases and incidence rate in Spain (2016–2024)

| Province | Number of cases | Incidence rate |
|---|---|---|
| Araba/Álava | 37 | 11.09 |
| Albacete | 45 | 11.64 |
| Alicante/Alacant | 110 | 5.85 |
| Almería | 12 | 1.64 |
| Ávila | 8 | 5.05 |
| Badajoz | 33 | 4.93 |
| Balears, Illes | 49 | 4.18 |
| Barcelona | 759 | 13.28 |
| Burgos | 36 | 10.11 |
| Cáceres | 12 | 3.08 |
| Cádiz | 88 | 7.06 |
| Castellón/Castelló | 48 | 8.18 |
| Ciudad Real | 21 | 4.26 |
| Córdoba | 19 | 2.45 |
| Coruña, A | 116 | 10.36 |
| Cuenca | 8 | 4.09 |
| Girona | 105 | 13.35 |
| Granada | 113 | 12.26 |
| Guadalajara | 6 | 2.26 |
| Guipuzkoa | 100 | 13.77 |
| Huelva | 69 | 13.12 |
| Huesca | 22 | 9.81 |
| Jaén | 23 | 3.67 |
| León | 46 | 10.18 |
| Lleida | 33 | 7.5 |
| Rioja, La | 35 | 10.94 |
| Lugo | 54 | 16.56 |
| Madrid | 725 | 10.74 |
| Málaga | 91 | 5.37 |
| Murcia | 64 | 4.21 |
| Navarra | 44 | 6.65 |
| Ourense | 27 | 8.85 |
| Asturias | 88 | 8.7 |
| Palencia | 16 | 10.06 |
| Palmas, Las | 46 | 4.08 |
| Pontevedra | 112 | 11.86 |
| Salamanca | 33 | 10.08 |
| Santa Cruz de Tenerife | 42 | 4.02 |
| Cantabria | 88 | 15.06 |
| Segovia | 14 | 9.11 |
| Sevilla | 334 | 17.15 |
| Soria | 4 | 4.51 |
| Tarragona | 78 | 9.49 |
| Teruel | 10 | 7.43 |
| Toledo | 18 | 2.54 |
| Valencia/València | 134 | 5.18 |
| Valladolid | 139 | 26.76 |
| Bizkaia | 123 | 10.66 |
| Zamora | 13 | 7.7 |
| Zaragoza | 72 | 7.44 |
| Ceuta | 0 | 0 |
| Melilla | 0 | 0 |

Incidence rates are expressed per 100,000 population.

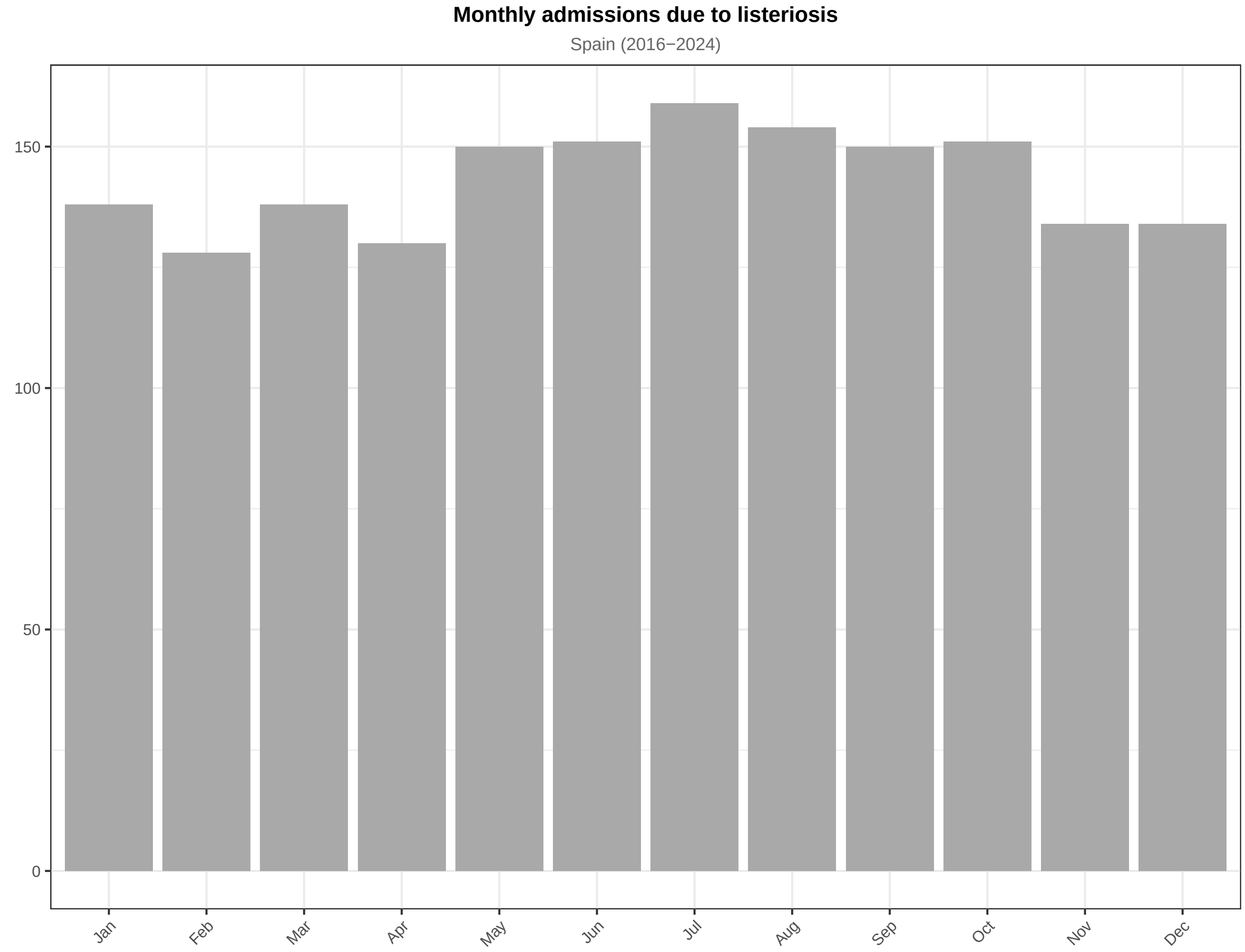


**Fig. S1.** Monthly cases of listeriosis during the observation period.

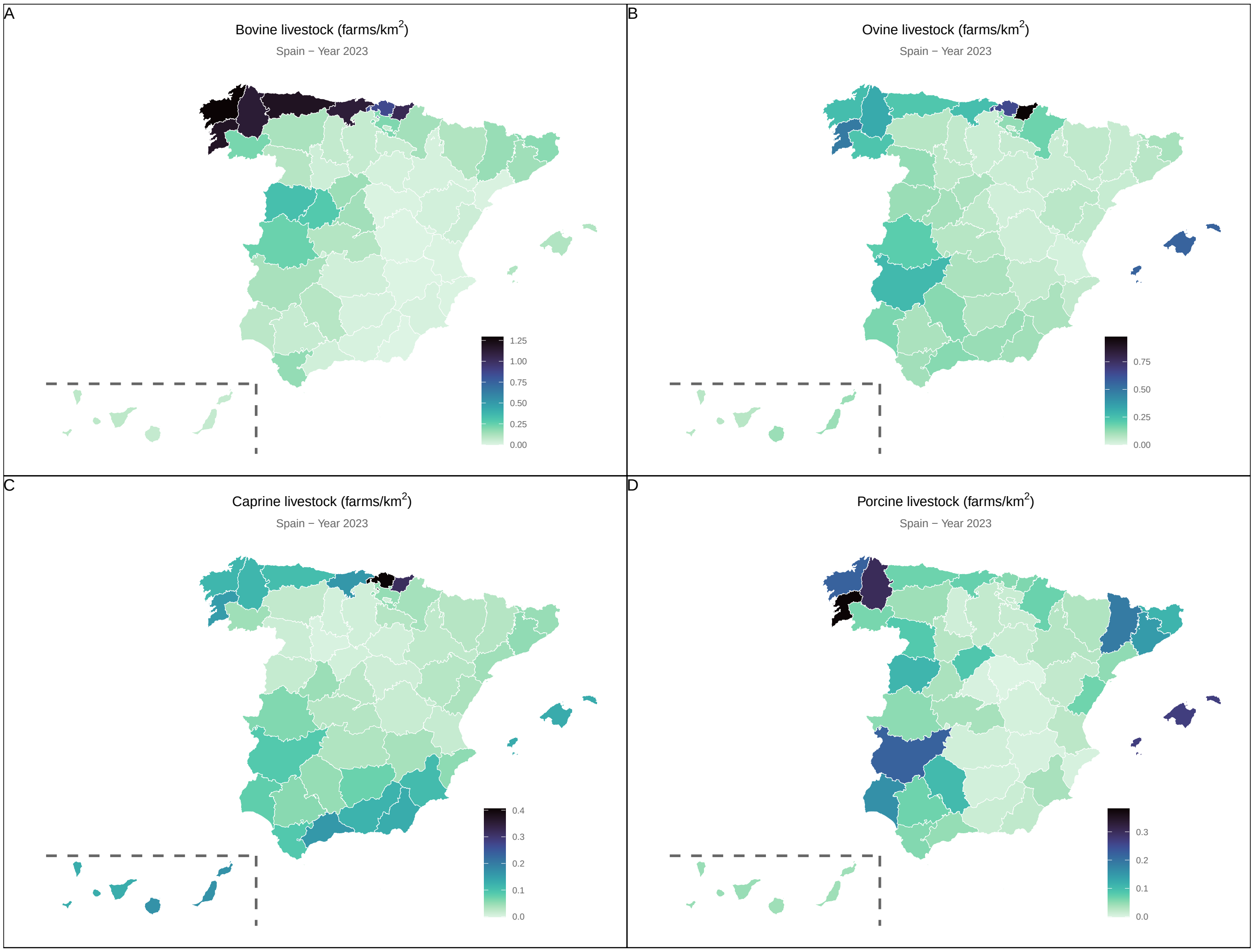


**Fig. S2.** National distribution of livestock, in density of farms/km$^2$.

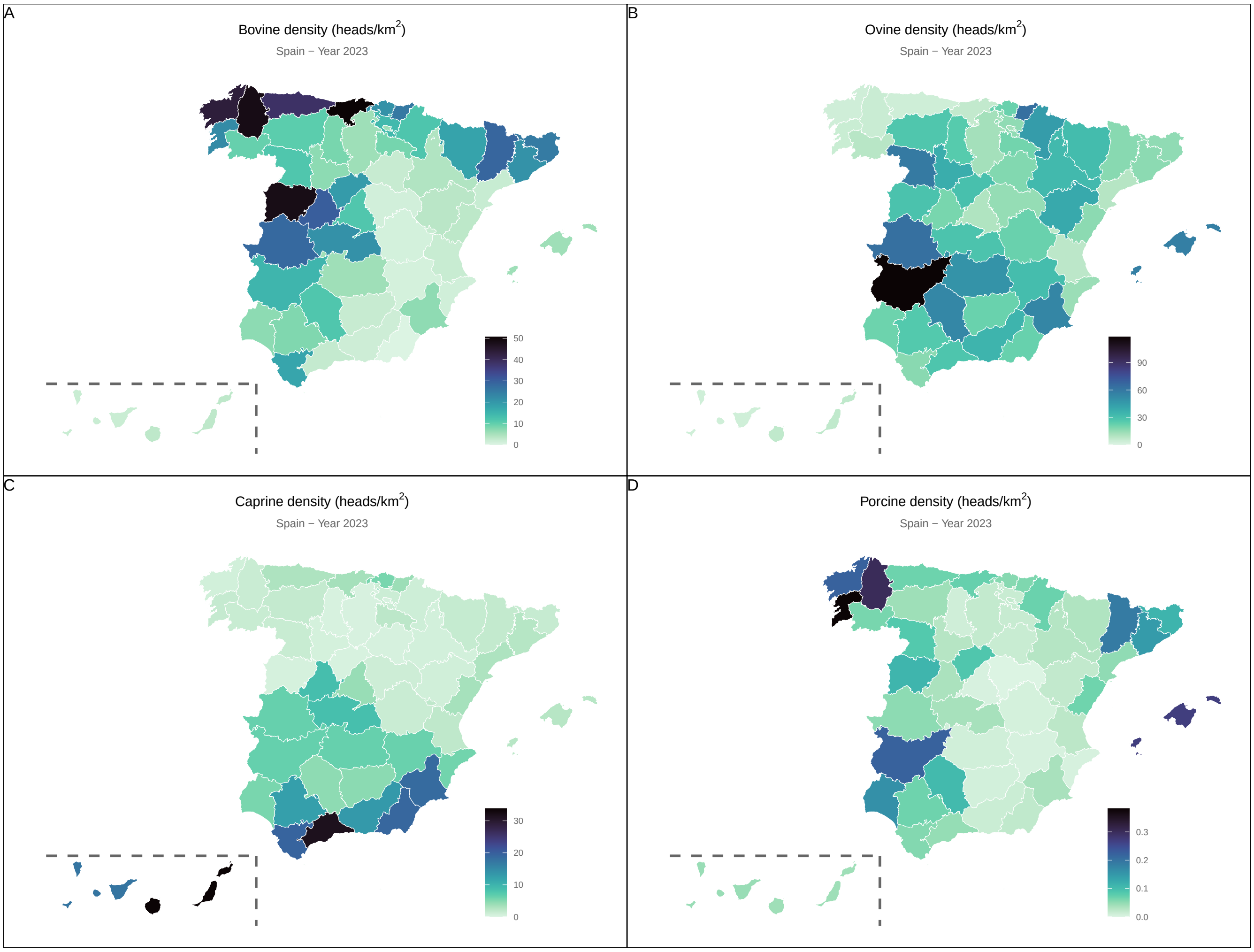


**Fig. S3.** Density of livestock, in heads/km$^2$.

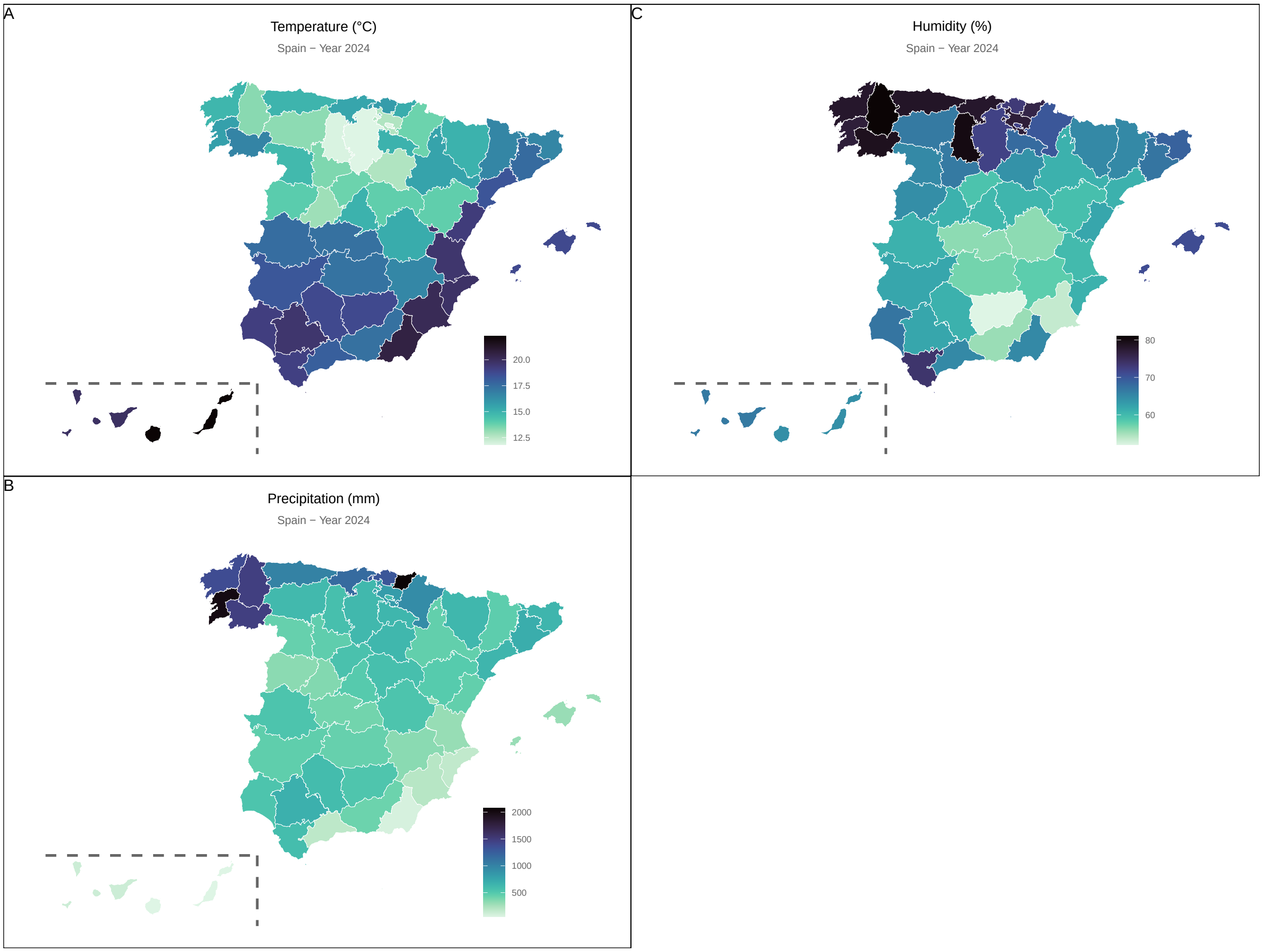


**Fig. S4.** Weather conditions in Spain (2024).
